# Developing multiplex ddPCR assays for SARS-CoV-2 detection based on probe mix and amplitude based multiplexing

**DOI:** 10.1101/2020.10.05.20207506

**Authors:** Raphael Nyaruaba, Changchang Li, Caroline Mwaliko, Matilu Mwau, Nelson Odiwour, Elishiba Muturi, Caroline Muema, Jin Xiong, Junhua Li, Junping Yu, Hongping Wei

**Author notes:** **Corresponding author:** Hongping Wei; Key Laboratory of Special Pathogens and Biosafety, Center for Biosafety Mega-Science, Wuhan Institute of Virology, Chinese Academy of Sciences, Xiao Hong Shan, #44 Bayi road, 430071, China.

## Abstract

Multiplexing has been highlighted to save on costs, increase sample throughput, and maximize on the number of targets that can be sensitively detected within a small sample. With the ongoing SARS-CoV-2 pandemic, different articles have been published highlighting the superiority of droplet digital PCR (ddPCR) over the gold reverse transcription PCR (RT-PCR) in SARS-CoV-2 detection. However, few studies have been reported on developing multiplex ddPCR assays for SARS-CoV-2 detection and their performance. In this study, we developed simplex (1 target), duplex (2 targets), triplex probe mix (3 targets), and fourplex (4 targets) assays based on a two color ddPCR system for SARS-CoV-2 detection. Results showed that the fourplex assay had the similar limits of detection and accuracy to the lower multiplex assays. Analyzing 94 clinical isolates demonstrated that the ddPCR triplex probe mix assay had better sensitivity than the RT-qPCR assay. Additionally, the ddPCR multiplex assay showed that remdesivir could inhibit the growth of SARS-CoV-2 *in vitro* while another testing drug couldn’t. Conclusively, our research shows that developing multiplex ddPCR assays is possible by combing probe mix and amplitude based multiplexing, which will help in developing multiplexed ddPCR assays for different SARS-CoV-2 applications.

## Introduction

Since the discovery and isolation of severe acute respiratory syndrome coronavirus 2 (SARS-CoV-2) in Wuhan, China [1], the virus has spread to multiple countries across the globe causing a global public health crisis. Many health systems have been crippled by this pandemic and all efforts are geared towards prevention, diagnosis, and treatment of the virus and its associated disease coronavirus disease 2019 (COVID-19).

As much as many companies and research institutes have come up with drugs, vaccines and other potent solutions to stop this virus, a permanent solution to stop this pandemic still doesn’t exist. This has left many countries to rely on diagnostics to detect, isolate, and treat infected patients. The most commonly used method and gold-standard for detection of infection cases in this pandemic is reverse transcription quantification real-time PCR (RT-PCR). PCR is not a new technique as it has grown since its discovery in the 1990’s [2] to become a well-established and commonly used technique to diagnose infectious diseases.

PCR has seen a transformation over the past decades since its discovery. The analogous qualitative conventional PCR that relies on gel electrophoresis was improved to quantitative real time PCR (qPCR) for detection and relative quantification. However, due to the need of a standard curve to perform relative quantification using qPCR, the third generation of PCR, digital PCR (dPCR), was developed to absolutely quantify pathogens directly [3]. Since its advent, dPCR has undergone lots of technological improvements and applications that are slowly overshadowing qPCR [4,5]. Technological refinements in dPCR have led to the development of better platforms like droplet digital PCR (ddPCR) capable of portioning samples to thousands of droplets for direct detection and quantification. Recent publications have highlighted the superiority of dPCR [3] in detection and quantification of targets including SARS-CoV-2 [6–13] when compared to RT-PCR. In their publication [7], Suo et al. showed that dPCR could detect SARS-CoV-2 reliably in clinical samples that tested negative by RT-PCR. This meant that the assay detection capabilities were superior to that of RT-PCR and it could avoid false negative results that were recorded by RT-PCR.

Similar to RT-PCR, multiplexing is possible by digital PCR [14]. Using probe based or Evagreen assays, one can design multiple assays to quantify more than one target in a single well. This has been highlighted to save on costs when running simplex assays and also increase the chances of target detection [15]. So far, no publication has been made publicly available to explain the different approaches and steps on how to develop multiplex ddPCR assays that can detect SARS-CoV-2. The available multiplexed ddPCR assays have been commercialized and don’t explain steps on how to develop similar assays. This poses a challenge to prospective researchers and clinicians who want to develop in-house multiplex assays for SARS-CoV-2 detection, diagnosis, and/or research. In this study, we used already ddPCR tested [6–9,11] China CDC SARS-CoV-2 primers [16] and additional in-house primers to show steps on how to develop simplex, duplex, triplex probe mix, and fourplex assays that can reliably detect and also quantify SARS-CoV-2 from clinical and research samples.

## Materials and methods

### Ethical Considerations

Wuhan Institute of Virology (WHIOV) is among the labs and institutes approved by China CDC of Wuhan city to conduct research on SARS-CoV-2 and detect COVID-19 from clinical samples. Research on developing new diagnostic techniques for COVID-19 using clinical samples has also been approved by the ethical committee of Wuhan Institute of Virology (2020FCA001).

During the outbreak in Wuhan, numerous clinical isolates from different hospitals were transported to WHIOV for detection. To ensure biosafety, all samples were received and inactivated first in the biosafety level 2 laboratory (BSL 2) with personal protection equipment for biosafety level 3 (BSL 3) laboratory.

### Sample categories for assay development

Since the assays may be used for research or diagnosis, three sample categories were used to develop the various ddPCR assays. Sample 1 was a sample containing only the SARS-CoV-2 genome (SARS-CoV-2 only) obtained from cultured virus in Vero E6 cells to represent research work; sample 2 was the human gene sample (IC only) pooled from oral swabs of healthy volunteers to represent negative clinical samples during diagnosis; and sample 3 was the SARS-CoV-2 virus from Vero E6 cells spiked with the human gene (SARS-CoV-2+IC) to represent a positive clinical sample during diagnosis.

### Sample processing

All the samples used in this work were first inactivated in the BSL 2 laboratory by heating at 56°C for 30 min. Inactivated samples were then extracted (bead extraction) using the Purifier™ Modesty automated RNA extraction machine. Post extraction, the RNA was converted to cDNA using the TakaRa PrimeScript™ RT Master Mix according to the manufacturer’s instructions for subsequent experiments (RT-qPCR and/or ddPCR).

### Primers and probes

China CDC Primers and probes (Table 1) targeting the open reading frame 1ab (ORF1ab) and nucleoprotein (N) gene regions [16] of SARS-CoV-2 were used to develop the various assays. Two additional primer sets targeting the receptor binding domain 2 (RBD2) of SARS-CoV-2 and an endogenous internal control Ribonuclease P protein subunit p30 (RPP30) [8] targeting the human gene were also synthesized and used to develop the various assays. All primers and probes were synthesized by the Sangon Biotech Company, Beijing, China.

**Table 1:**
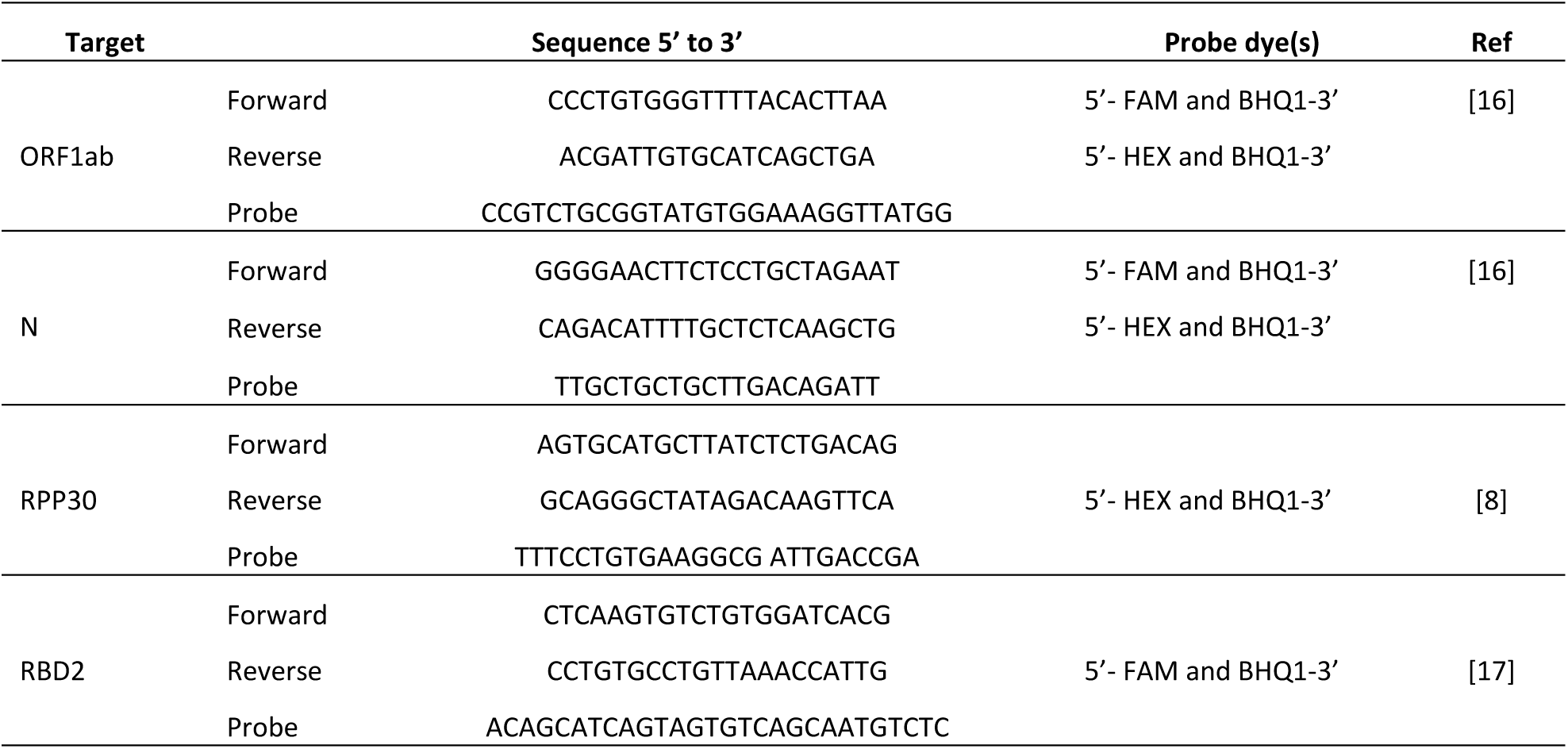
Primer and probe sequences for SARS-CoV-2 and an endogenous human control gene.

### Real-Time PCR Assay Composition

The Luna^®^ Universal probe qPCR Master Mix (New England BioLabs) was used for developing the qPCR assay according to the manufacturer’s instructions. The qPCR assay was composed of 0.4 µM and 0.2 µM primer and probe (table 1) concentrations respectively, 10 µL Luna Universal qPCR Probe Master Mix, 2 µL cDNA sample, and nuclease free water to a final volume of 20 µL. All qPCR experiments were performed in a CFX96 Touch™ real-time PCR (BioRad) instrument under the reaction conditions initial denaturation at 95 °C for 5 sec, followed by 40 cycles of denaturation at 95 °C for 5 sec and annealing (reading step) at 64 °C for 1 minute.

### Droplet digital PCR workflow and assay composition

The ddPCR workflow begins with making the detection assay mix. The ddPCR™ Supermix for Probes (No dUTP) was used to develop all the assays after generating cDNA. Different primer and probe concentrations were used to develop the simplex, duplex, triplex probe mix, and fourplex assays. The 1× concentration of the various assays included:

#### Simplex assays

Each assay here was composed of a primer and probe pair for a particular target. The general 1x assay was composed of 11 µL 2× ddPCR Supermix for probes (No dUTP), 900 and 250 nM primer and probe concentrations respectively, 2.2 µL cDNA, and nuclease free water to a final volume of 22 µL.

#### Duplex assays

The duplex assay was composed of two sets of primers and probes. Each duplex assay was set to detect a target (N/ORF1ab) in the FAM channel and the human gene (IC) RPP30 in the HEX channel. The 1× assay was similar to that of the simplex assay with only a variation in the volume of nuclease free water due to the addition of a new primer set.

#### Triplex probe mix assay

The triplex probe mix assay was composed of three sets of primers and probes to detect two targets (ORF1ab and N) and the IC (RPP30). The general 1× assay was composed of 11 µL 2× ddPCR Supermix for probes (No dUTP), 900 and 250 nM primer and probe concentrations respectively, 2.2 µL cDNA, and nuclease free water to a final volume of 22 µL. However, for the probes, the targets were labelled with different dyes (FAM and/or HEX) at different ratios. Target 1 was labelled with FAM (1:0), target 2 was labelled with 50% FAM and 50% HEX (1:1), and target 3 was labelled with HEX (0:1). Target 1 and 2 were ORF1ab and N gene used interchangeably while target 3 was RPP30.

#### Fourplex amplitude based assay

The fourplex assay was composed of four sets of primers and probes to detect three SARS-CoV-2 targets (ORF1ab, N, and RBD2) and the IC (RPP30). The 1× fourplex assay was composed of 11 µL 2× ddPCR Supermix for probes (No dUTP), primers at 900 nM (1×) and 450 nM (0.5×) concentrations, probes at 250 nM (1×) and 125 nM (0.5×) concentrations, 2.2 µL cDNA, and nuclease free water to a final volume of 22 µL. Each cannel was used to detect two targets at 0.5× and 1× primer and probe concentrations. For FAM targets, RBD2 primers and probe were added at a concentration of 0.5× while N primers and probe were added at 1× concentrations. For the HEX channel, RPP30 primers and probe were added at 0.5× concentrations while ORF1ab primers and probe were added at 1× concentrations. Since this assay included an in-house primer set (RBD2), it was excluded in subsequent optimization experiments and only used for demonstration purposes.

After distributing the different assays in a ddPCR plate, approximately 20,000 nanoliter-sized droplets per well were generated using an automated droplet generator (QX200™ AutoDG ddPCR system (Bio-Rad, US)). Resultant droplets were then heat-sealed with a pierceable aluminum foil using a PX1 PCR plate sealer (Bio-Rad) set to run at 180 °C for 5 sec before being loaded into a C100 Touch™ Thermal Cycler (Bio-Rad, US) for amplification. Thermal cycling with a ramp rate of 2°C/sec at every step was set to run for 10 min enzyme activation at 95 °C; followed by 40 cycles of denaturation at 94°C for 30 sec and 1 min annealing/extension at 57°C; enzyme deactivation at 98°C for 10 min; and a final hold step at 4°C for at least 30 minutes to stabilize the droplets. Post amplification, droplets were read in a QX200™ Droplet.

### Annealing temperature optimization

During thermal cycling of the droplets, a temperature gradient between 65°C to 55°C was inserted at the annealing temperature step to determine the best temperature for which droplet separation will be achieved in the simplex, duplex, and triplex probe mix assays.

### Inter and intra assay variability

Eight replicates per assay of a positive sample spiked with a human gene were used to test inter and intra assay variability. The sample was tested on simplex (ORF1ab, N, and RPP30 (IC), duplex (N and IC, and OFR1ab and IC), and triplex probe mix assays. For the triplex assays, two assays with interchanging N or ORF1ab targets to 50% (FAM and HEX) probe concentrations were used.

### Determining LoB/LoD

The triplex probe mix assay was used to determine SARS-CoV-2 detection limits. For Limit of Blank (LoB), 21 SARS-CoV-2 negative human samples and 3 non template controls (NTC) were used. However, due to the absence of a SARS-CoV-2 standard, SARS-CoV-2 cDNA from Vero E6 cells was serially diluted in a background of already pooled human oral swab matrix to determine the assay Limit of Detection (LoD). A total of 16 two-fold serial dilutions were done and detected in triplicates using the triplex probe assay. The LoD was set to be the lowest concentration at which two of the three (2/3) triplicate samples could be detected in the dilution series. The LoD for individual targets (ORF1ab and N) were determined in copies/µl and copies/PCR.

### Application of the multiplex assay

The triplex probe mix assay approach was used to demonstrate the applicability of multiplex assays in detecting clinical samples and performing research. For the clinical assay, 94 clinical samples that were negative or with low virus concentrations were used to test the triplex probe mix ddPCR assay (N (1:0), ORF (1:1), and RPP30 (0:1)). The same primers were also used to perform qPCR. A one-step commercial RT-qPCR kit (DaAn Gene Co., Shenzhen) targeting the ORF1ab (VIC), N (FAM), and an endogenous internal control (Cy5) was used according to the manufactures instructions to validate the qPCR and ddPCR detection results.

For the research application, we tested the applicability of the triplex probe mix assay (RBD2 (1:0), N (1:1), and ORF1ab (0:1)) in determining the inhibition efficiency of remdesivir and a drug we named Code 30 (due to ongoing research) against SARS-CoV-2 *in vitro*. Vero E6 cells were infected with SARS-CoV-2 at a multiplicity of infection (MOI) of 0.001. One set was used as a control while the other two sets were infected with remdesivir and Code 30 to a final concentration of 10 µM. 24 hours post infection (hpi), 200 µL of the cell supernatant from all sets was extracted in triplicate and the RNA quantified using the ddPCR triplex probe mix assay.

### Data analysis

A droplet count of ≥ 10,000 droplets was set as the cutoff when analyzing all ddPCR experiments [15]. If the number of droplets were lower, the results were discarded or the experiment repeated. All ddPCR data were generated using BioRad’s QuantaSoft™ software version 1.7.4.0197. Simplex and duplex assay experiments were analyzed directly using the same software. However, for the triplex probe mix and fourplex assays, the QuantaSoft™ Analysis Pro software version 1.0.596 was used for analysis. Further statistical analysis was done using the GraphPad Prism Version 6.01 software.

## Results

### Droplet separation

Three sample categories highlighted in the methodology section were used to determine how droplets separated in the simplex, duplex, triplex probe mix, and fourplex assays. Since it is important to have a control during diagnosis, the RPP30 gene was included in all multiplex assays. The results were as follows:

#### Simplex assay

Droplets could be detected in all the simplex assays as seen in Figure 1. Both the ORF1ab and N gene targets were labelled with FAM hence read in Channel 1. From the results, the virus could be detected in the sample categories with only the virus (V) and the virus spiked in a background of pooled human gene (V+H). No droplets were observed for both the ORF1ab and N genes when only the healthy human sample (H) and negative (Neg) control samples were detected as seen in Figure 1A and B. The RPP30 was labelled with HEX hence read in Channel 2. The human gene could be detected in only the H, and V+H samples as seen in Figure 1C. The human gene could not be detected in the V and Neg samples as they didn’t contain the human gene.

**Figure 1:**
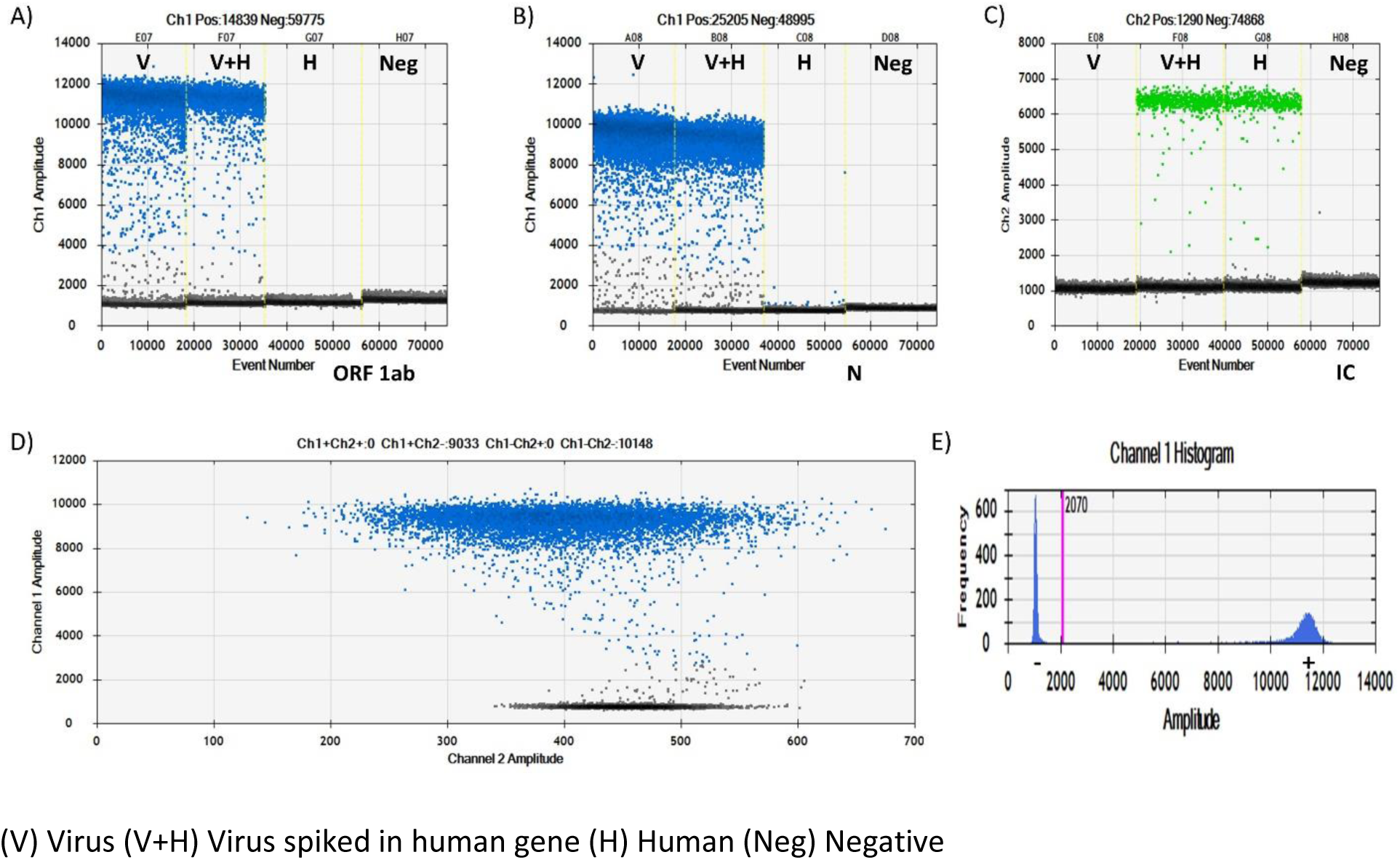
Simplex assay droplet separation results. A,B, and C are 1D amplitude results of the ORF1ab, N, and RPP30 targets respectively using various sample categories. D is a representative 2D amplitude result. E is a representative histogram result of a simplex assay.

#### Duplex assay

For the duplex assays, one target was FAM labelled (ORF1ab) and the other HEX labelled (RPP30). This meant that droplets were to be observed in both Channel 1 and Channel 2 as seen in Figure 2. As expected, four droplet clusters were observed in the 2D channel and two in the 1D channel (for each target) when a sample containing the virus spiked in a background of pooled human gene was detected (Figure 2B). However, when a sample contained only the virus was read (Figure 2A), we observed a positive signal in the FAM channel and no signal in the HEX channel with a consequent 2 droplet amplitude partition in the 2D channel. The reverse was true when only a sample containing only the human gene was detected (Figure 2C).

**Figure 2:**
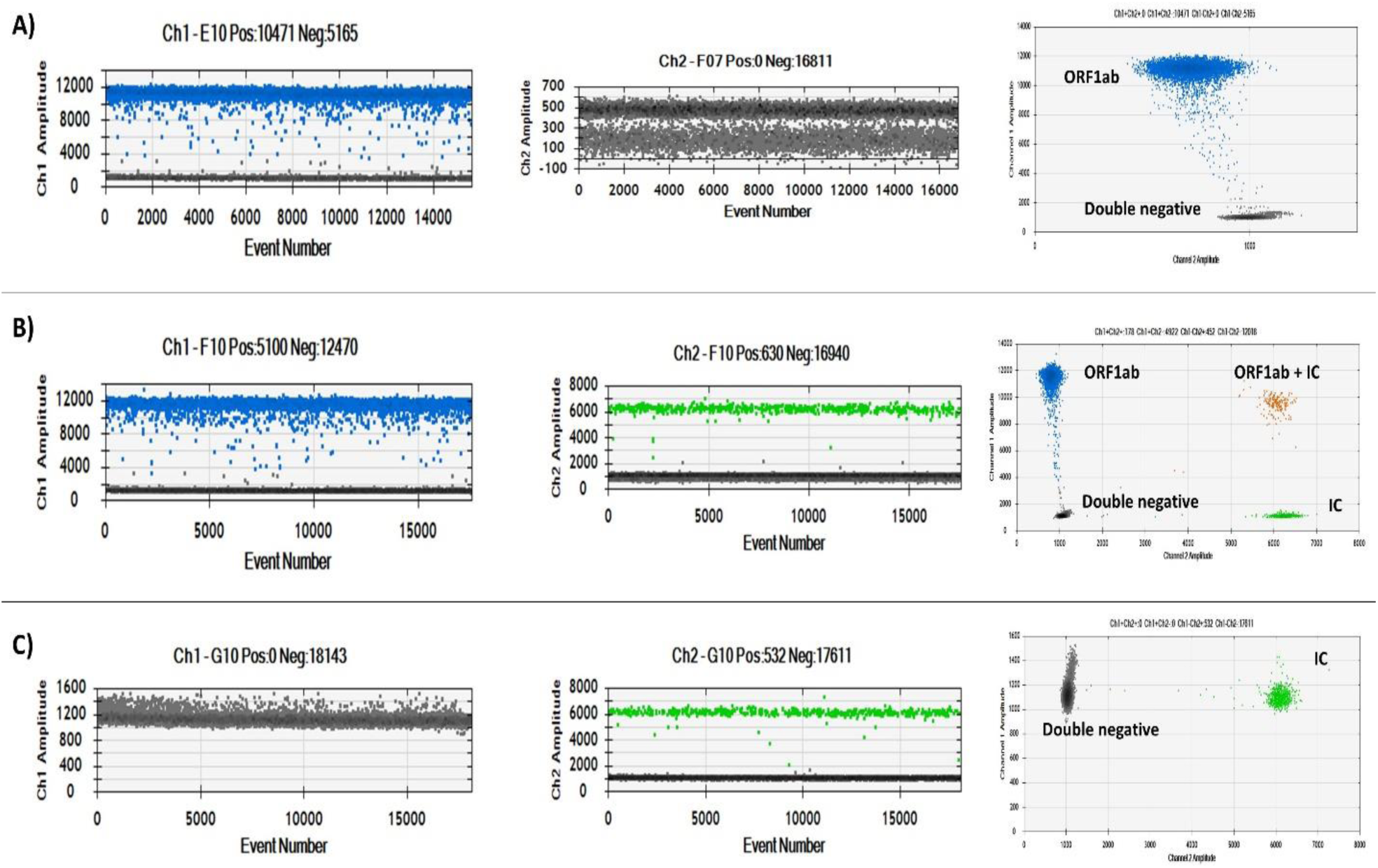
Duplex assay droplet separation results in the 1D and 2D amplitudes. A) SARS-CoV-2 only sample. B) SARS-CoV-2+IC sample. C) IC only sample.

#### Triplex probe mix assay

Eight droplet clusters were observed (Figure 3A) when a triplex probe mix triplex assay was run to detect SARS-CoV-2 spiked in a background of pooled human sample. The normal QuantaSoft™ software used for reading droplets could not classify which targets the droplets belonged to as seen in Supplementary Figure S1. To analyze the data, we used the QuantaSoft™ Analysis Pro software as it was designed to curb this inefficiency. Using the circular threshold tool in the software, the droplet clusters were designated as follows in the 2D amplitude: triple negative (bottom left gray cluster); N (single positive (FAM channel) directly above the triple negative)); ORF1ab (single positive (FAM and HEX channels) at a ∼45° angle from the triple negative)); IC (single positive (HEX channel) immediately to the right of triple negative)); N+ORF1ab (double positive above N and ORF1ab); N+IC (double positive above IC); ORF1ab+IC (double positive at a 45° angle from IC); and N+ORF1ab+IC (triple positive and right above ORF1ab+IC).

**Figure 3:**
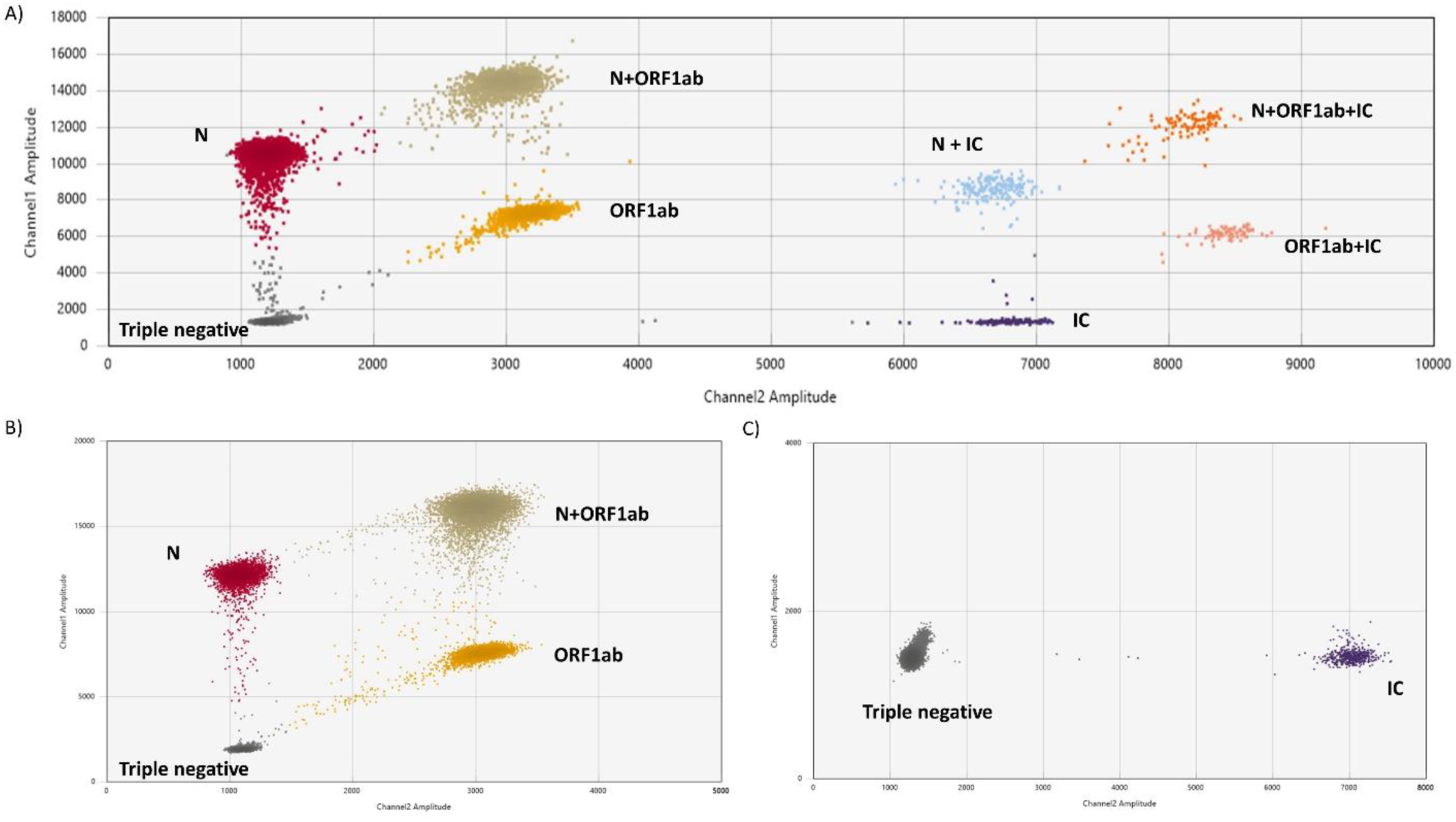
Droplet separation in a triplex probe mix assay results. A) SARS-CoV-2+IC sample. B) SARS-CoV-2 only sample. C) IC only sample.

From the different sample categories in the triplex probe mix assay, the clusters of double positives and triple positive reduce when only the virus (Figure 3B) or the human gene (Figure 3C) is detected. Additionally, the target cluster positions are maintained with different samples.

#### Fourplex amplitude based assay

Sixteen droplet clusters were observed in the fourplex assay using a sample containing SARS-CoV-2 spiked in a background of pooled human sample. Similar to the triplex assay, the QuantaSoft™ software used for reading the droplets could not directly classify the clusters as seen in Figure S2, hence, the QuantaSoft™ Analysis Pro software was used. The droplet clusters were assigned using the circular threshold tool available in the QuantaSoft™ Analysis Pro software. The droplets were clustered as: quadruple negative (bottom left gray cluster); RBD2 (single positive (FAM channel) directly above the quadruple negative)); N (single positive (FAM channel) directly above RBD2)); RBD2+N (double positive (FAM channel) directly above N)), IC (single positive (HEX channel) immediately to the right of quadruple negative)); ORF1ab (single positive (HEX channel) immediately to the right of IC)); IC+ORF1ab (double positive (HEX channel) immediately to the right of ORF1ab)); and quadruple negative (highest droplet cluster above IC+ORF1ab)). Resultant double positive and triple positive droplets clusters of different targets could be located at the intersections of the arrows. The different sample types also helped in the location of different clusters. From the amplitude result of the 2D channel, the droplet clusters of the SARS-CoV-2 targets were maintained at the same positions even when different sample categories were used (Figure 4).

**Figure 4:**
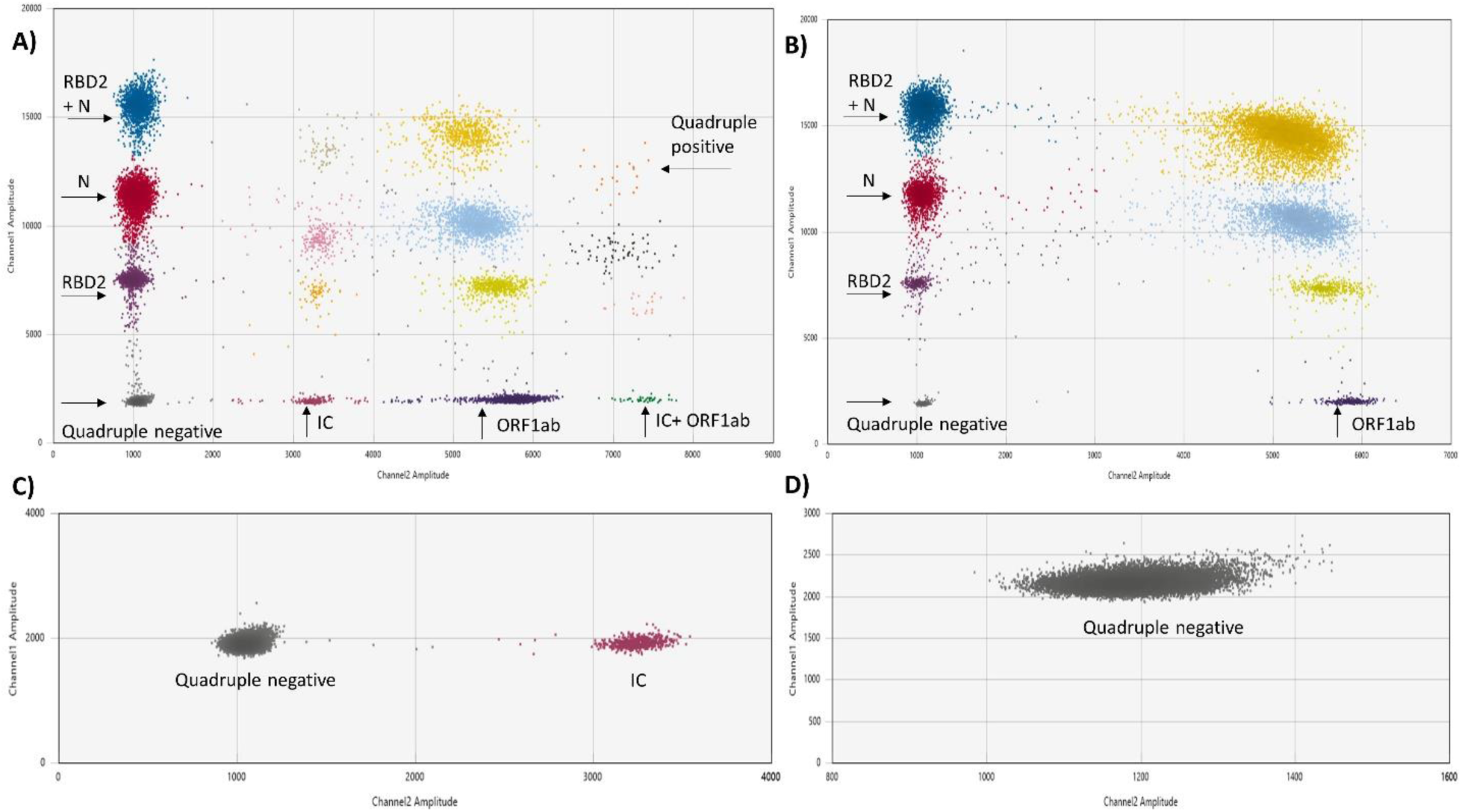
Droplet separation in a fourplex amplitude based assay. A) SARS-CoV-2 spiked in a background of pooled human sample. B) SARS-CoV-2 sample. C) Pooled human sample. D) Negative sample.

### Temperature gradient analysis

As seen in Figure 5, there was a general increase in the positive droplet amplitude with a decrease in annealing temperature from 65°C to 55°C. In the 2D channel of both the duplex and triplex probe mix assays, there was no clear separation of droplet clusters till the temperatures dropped from 65°C to 63°C. Even though clusters could be assigned at 63°C, better separation was yet to be achieved. For the duplex assay, the 1D plot results (data not shown) were similar those of the simplex assays. An annealing temperature of 57°C was found optimum for subsequent experiments as optimum separation was achieved at 56.9°C. This temperature was suitable for all the assays.

**Figure 5:**
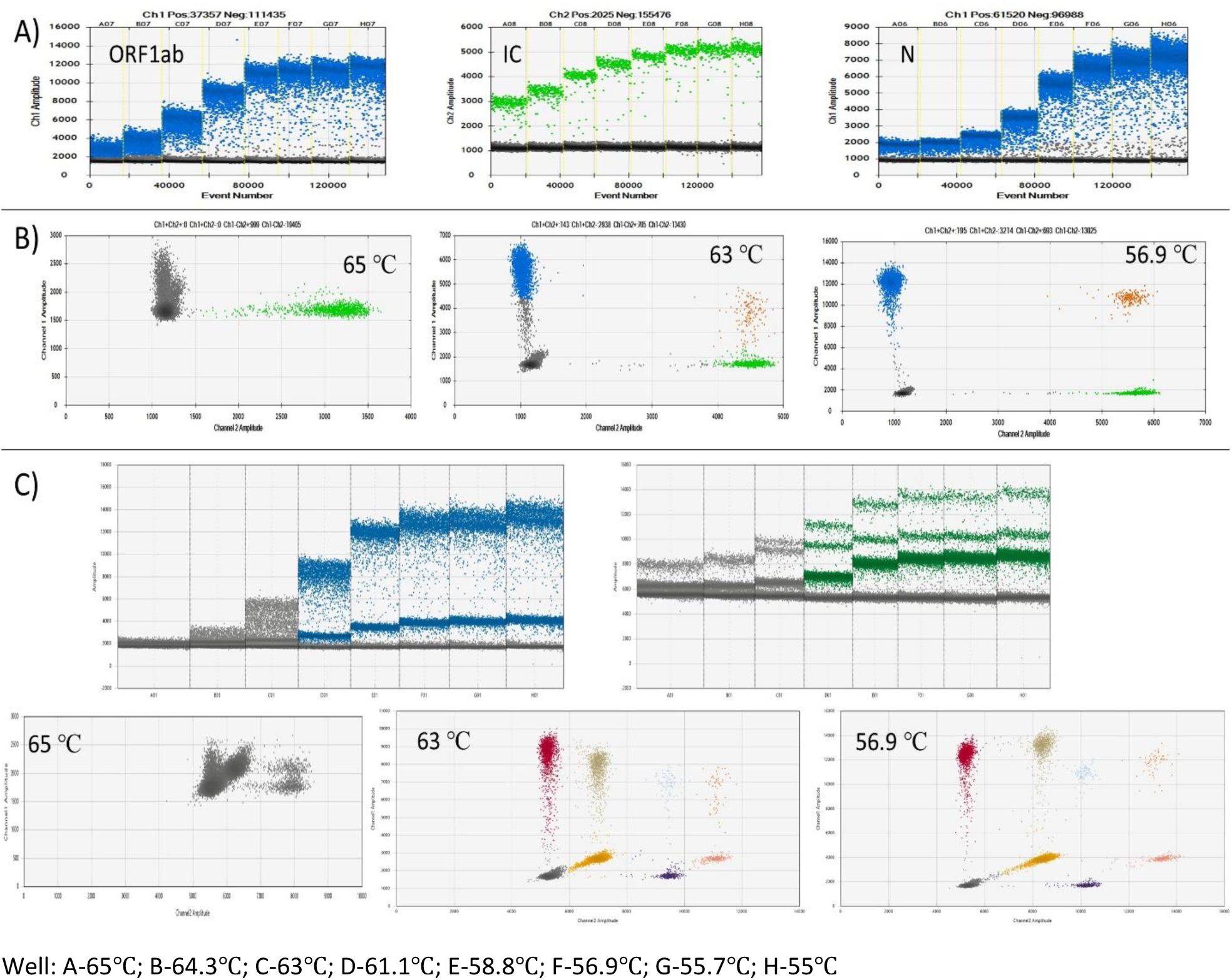
Temperature gradient analysis of simplex (A), duplex (B), and triplex probe mix (C) assays from 65°C to 55°C. There was a general increase in droplet amplitude with a decrease in annealing temperature. Optimum separation was achieved at an annealing temperature of 56.9°C in all the assays.

### Assay variability

Intra- and inter-assay variability of replicate wells was used to determine reproducibility of results using different assays. The percentage co-efficient of variation (%CV) of all tested assays was determined. As seen in Table 2, there was little intra- and inter-assay variability (%CV < 6) between replicate wells of simplex, duplex, and triplex probe mix assays post quantification.

**Table 2:**
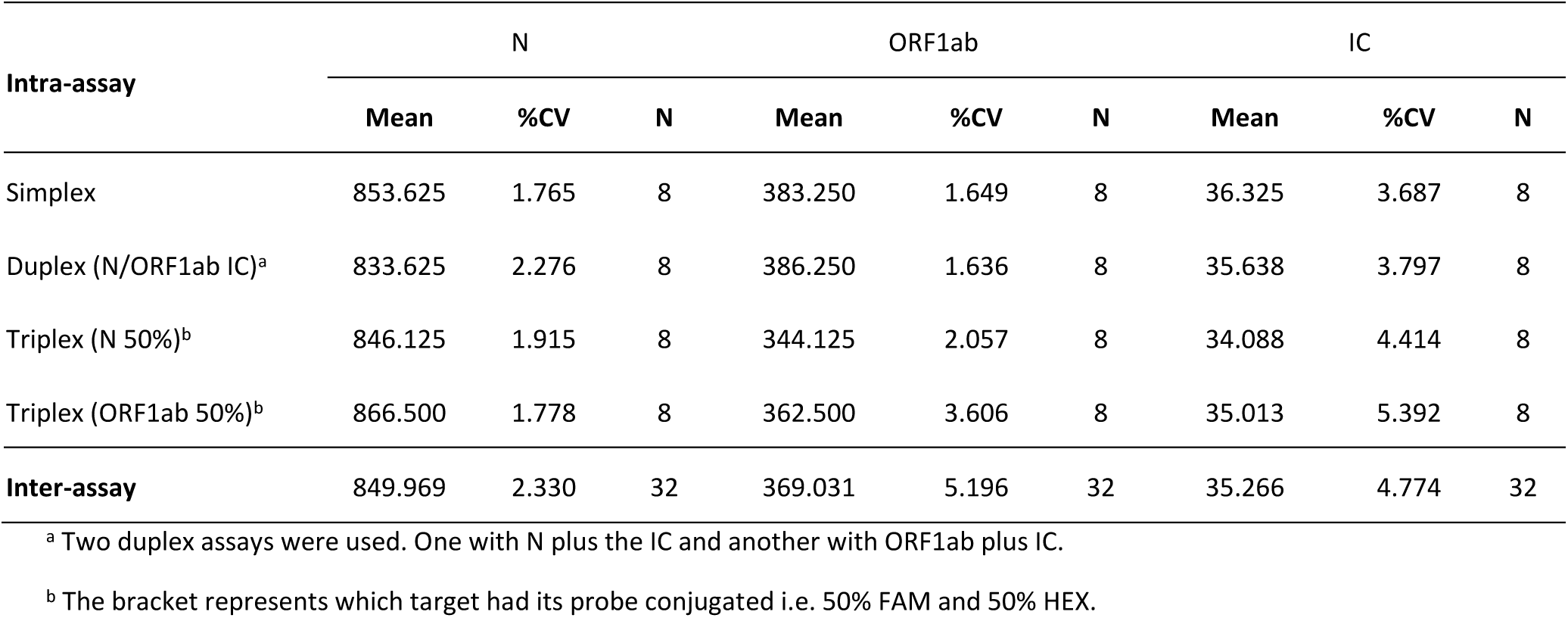
Inter- and intra-assay variability of replicate wells.

| Intra-assay | N |  |  | ORF1ab |  |  | IC |  |  |
| --- | --- | --- | --- | --- | --- | --- | --- | --- | --- |
|  | Mean | %CV | N | Mean | %CV | N | Mean | %CV | N |
| Simplex | 853.625 | 1.765 | 8 | 383.250 | 1.649 | 8 | 36.325 | 3.687 | 8 |
| Duplex (N/ORF1ab IC)<sup>a</sup> | 833.625 | 2.276 | 8 | 386.250 | 1.636 | 8 | 35.638 | 3.797 | 8 |
| Triplex (N 50%)<sup>b</sup> | 846.125 | 1.915 | 8 | 344.125 | 2.057 | 8 | 34.088 | 4.414 | 8 |
| Triplex (ORF1ab 50%)<sup>b</sup> | 866.500 | 1.778 | 8 | 362.500 | 3.606 | 8 | 35.013 | 5.392 | 8 |
| **Inter-assay** | 849.969 | 2.330 | 32 | 369.031 | 5.196 | 32 | 35.266 | 4.774 | 32 |

<sup>a</sup> Two duplex assays were used. One with N plus the IC and another with ORF1ab plus IC.
<sup>b</sup> The bracket represents which target had its probe conjugated i.e. 50% FAM and 50% HEX.

All the assays also had high reproducibility as target copies were almost at par as seen in Figure 6. This meant the triplex probe mix assay performed similarly to the simplex and duplex assay hence found fit for further experiments.

**Figure 6:**
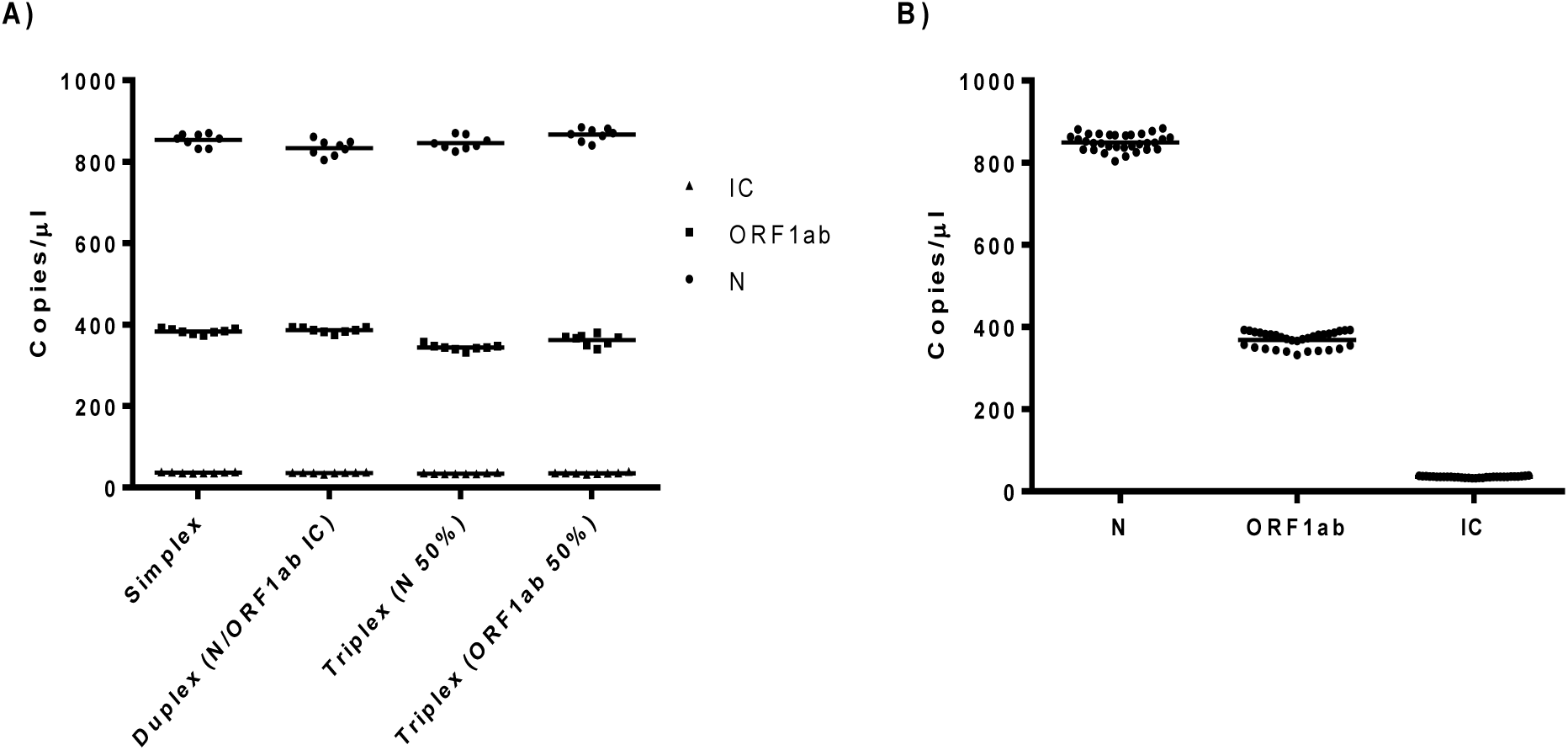
Reproducibility of replicate wells. A) Intra assay reproducibility of eight replicate wells using different assays. B) General reproducibility after the replicate wells data of each target in all assays are merged together.

### Assay LoB and LoD

From the 24 samples used to determine the triplex probe mix assays LoB, 4/24 samples had one droplet in the N target with 3/4 arising from human samples, and 1/4 from NTC samples as seen in **Table S1**. Hence, the LoB for the N target and ORF1ab target was determined to be 1 and O positive droplets respectively. After detection of the serially diluted cDNA, the LoD for N and ORF1ab target was determined to be 0.12 copies/µL (2.64 copies/PCR) and 0.13 copies/µL (2.86 copies/PCR), respectively. Consequently, a threshold of at least two droplets in either or both targets was concluded for classifying a sample to be SARS-CoV-2 ddPCR positive.

### Fitness for purpose

The triplex probe mix assay was used to investigate the fitness of the multiplex assay in diagnosis and research.

#### Diagnosis

94 clinical isolates were used to test the performance of the triplex probe mix assay in diagnosis of COVID-19. The same targets used in the triplex probe mix assay were also detected using qPCR. Both the two assays were validated using the DaAn commercial RT-qPCR kit for SARS-CoV-2 detection in clinical samples. As summarized in table 3, the ddPCR triplex probe mix assay performed better than the qPCR assay.

**Table 3:** Diagnostic performance of the triplex probe mix and qPCR assays in detecting clinical samples.

| N=94 |  | Commercial qPCR kit |  | Sensitivity<br>(95% CI) | Specificity<br>(95% CI) | PPV<br>(95% CI) | NPV<br>(95% CI) | NLR<br>(95% CI) | Accuracy<br>(95% CI) |
| --- | --- | --- | --- | --- | --- | --- | --- | --- | --- |
|  |  | Positive | Negative |  |  |  |  |  |  |
| ddPCR assay | Positive | 52 | 0 | 96.3% | 100% | 100% | 95.24% | 0.04 | 97.87% |
|  | Negative | 2 | 40 | (87.25 -99.55%) | (91.19-100%) | (N/A) | (83.7-98.73%) | (0.01-0.14) | (92.52-99.74%) |
| qPCR assay | Positive | 50 | 0 | 92.59% | 100% | 100% | 90.91% | 0.07 | 95.74% |
|  | Negative | 4 | 40 | (82.11-97.94%) | (91.19-100%) | (N/A) | (79.57-96.25%) | (0.03-0.19) | (89.46-98.83%) |

N-total samples; PPV-positive predictive value; NPV-negative predictive value; NLR-negative likelihood ratio
Calculated online by MEDCALC (https://www.medcalc.org/calc/diagnostic_test.php)

Notably, the sensitivity of the ddPCR assay was slightly higher than that of the qPCR as two samples that failed to be detected by qPCR were detected by both ddPCR and the commercial kit. This consequently improved the accuracy of ddPCR in detecting SARS-CoV-2 from COVID-19 samples when compared to qPCR.

#### Research

Two drugs remdsevir and Code 30 were used to demonstrate the fitness of the triplex probe mix assay (N (1:0), ORF1ab (1:1) and RBD2 (0:1)) in performing research. Of note, the RPP30 gene was excluded here as this research was done on Vero E6 cells. As seen in the Figure 7A, no growth occurred in Vero E6 cells infected with remdesivir 24 hours post infection (hpi). However, Code 30 grew equally as the control 24 hpi. This meant that Code 30 had no inhibitory effect against SARS-CoV-2 while remdesivir had the ability to inhibit SARS-CoV-2 *in vitro* as seen in figure 7B. At a concentration of 10 µM, remdesivir’s percentage inhibition efficiency against SARS-CoV-2 *in vitro* was calculated to be 99.53%, 98.90%, and 98.92% for targets N, ORF1ab, and RBD2, respectively.

**Figure 7:**
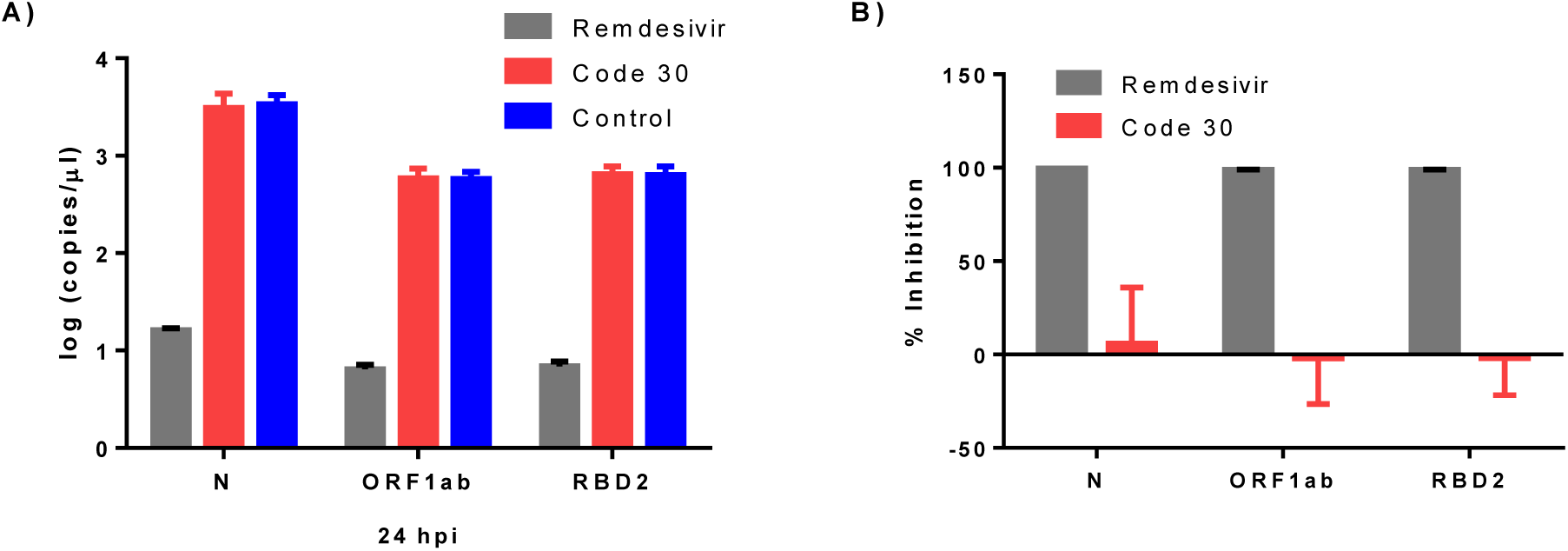
The antiviral activity of remdesivir and Code 30 against SARS-CoV-2 in cell culture. A) Quantified log copies/µL of different targets by ddPCR triplex probe mix assay 24 hpi. B) Inhibition efficiency of remdesivir and Code 30 against SARS-CoV-2 growth in Vero E6 cells.

## Discussion

Multiplexing has been used widely to save on costs, increase sample throughput, and maximize on the number of targets that can be sensitively detected within a small sample. When developing multiplexed qPCR assays, each target has to be labelled with a unique probe to differentiate the targets. In ddPCR, few systems have this ability to detect targets using more than two probes [14]. Detection in these systems is done in two discrete optical channels e.g. FAM and HEX/VIC as seen in Bio-Rads’ QX200 system that was used in this work. Despite this, ddPCR systems have a unique advantage that unlike qPCR, more than two targets can still be detected in a single reaction using higher order multiplexing. When developing higher order multiplex ddPCR assays, two main options exist: probe mix multiplexing and amplitude based multiplexing. In this article, we use already established primers that target different regions of the SARS-CoV-2 sequence to explain the two. To make it easy and help in the discussion, we developed a flowchart illustration **(Figure 8)** for reference on steps taken towards developing the multiplex assays for SARS-CoV-2 detection.

**Figure 8:**
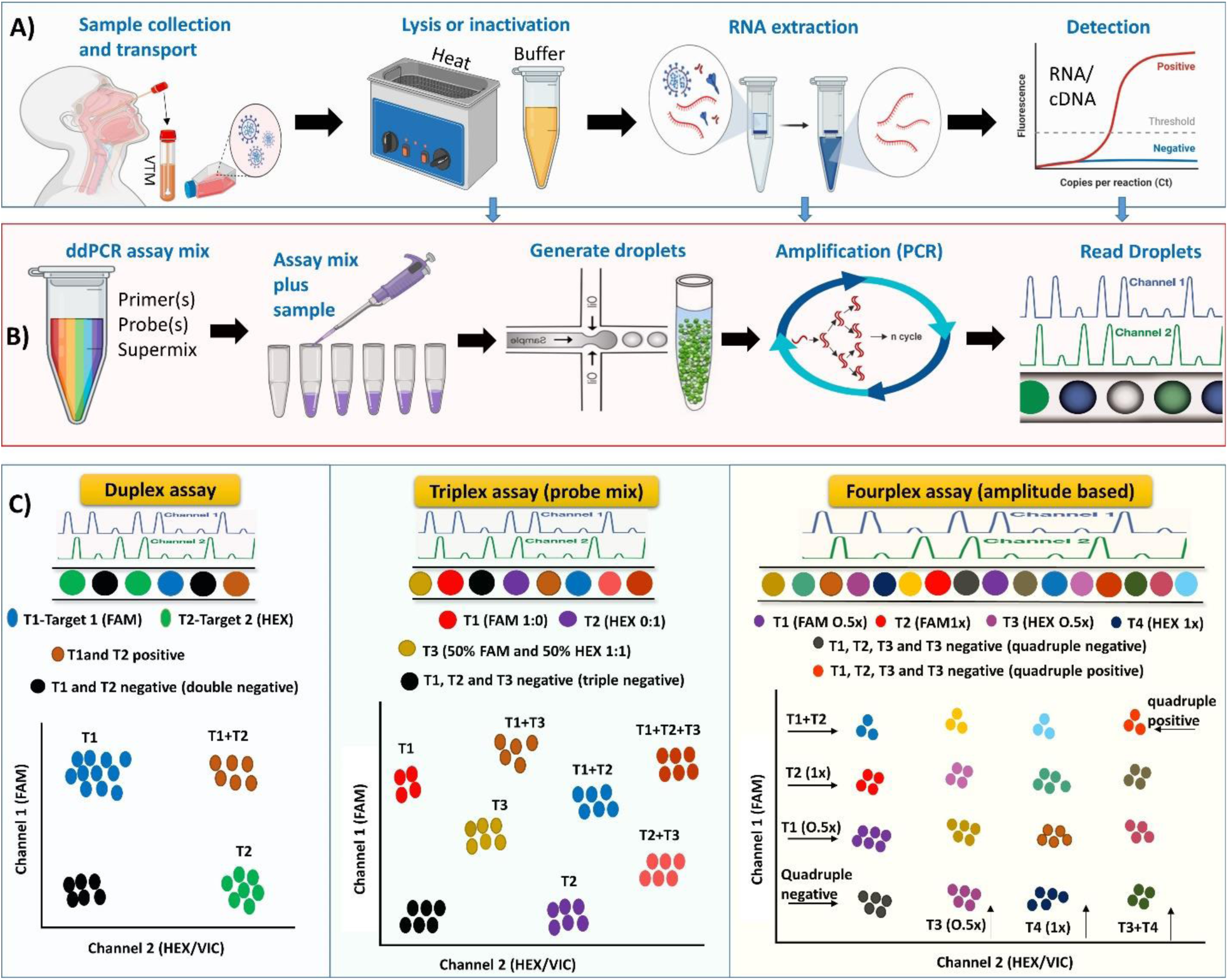
Flowchart for this research showing steps towards developing multiplex ddPCR assays (including higher order multiplex assays) for SARS-CoV-2. A) Sample handling and preparation workflow. B) ddPCR workflow. C) Data analysis and droplet separation.

Sample handling and preparation workflow: So far, clinical [6–8,10,12,13] and environmental [9,11] samples have been used for SARS-CoV-2 detection using ddPCR. Due to these advancements, we designed our workflow to not only aim at developing the multiplex assay for clinical application but also for research as seen in of Figure 8A. To actualize this, we developed three sample matrix containing only the virus (SARS-CoV-2) to represent laboratory research work, virus in a background of pooled human oral swabs (SARS-CoV-2 + IC) to represent clinical work, and a sample with only pooled human oral swabs (IC) negative for SARS-CoV-2 as a control representing clinical patients negative for SARS-CoV-2. Additionally, the three sample categories were used to help locate targets and assign droplet clusters when developing the assays.

Using ddPCR, it has been shown that SARS-CoV-2 could reliably be detected from nasopharyngeal samples without RNA extraction [18,19]. In our flowchart (Figure 8A), we also highlight this as a possibility even though we did not explore it. However, majority of the research done on SARS-CoV-2 using ddPCR have used samples that have undergone RNA extraction [6–13]. This is similar to our work. Due to the high costs incurred when performing ddPCR, we also recommended that SARS-CoV-2 samples can be detected by RT-qPCR first before ddPCR (last step in Figure 8A). As highlighted by Liu et.al. in their article [6], RT-qPCR can be used in mass diagnosis of SARS-CoV-2 due to low costs, however, in cases where sensitivity and precision are low, ddPCR would be the ideal method for quantification.

ddPCR workflow: The general ddPCR workflow is summarized in Figure 8B. The workflow begins with preparing your assay mix (primers/probe(s) mixed with the ddPCR supermix at different concentrations to make simplex or multiplex assays)). The assay mix and sample(s) are then added to PCR strips or 96-well PCR plates dependent on the ddPCR system. Post sample addition, droplets are generated using a droplet generator. Partitioning samples to generate thousands of droplets makes multiplexing by ddPPCR easy [14]. The droplets are then thermocycled to end-point. End-point detection post PCR helps ddPCR to become resistant to a number of efficiency differences between targets, improving on accuracy and precision [20]. Post amplification the droplets are read using a droplet reader to generate data for analysis.

Data analysis and droplet separation: As seen in Figure 8C, different approaches can be made to develop multiplex SARS-CoV-2 ddPCR assays. Dependent on the assay, droplets will be separated differently according to the florescence amplitude exhibited by the droplets. The most basic in this work was development of the duplex assays. Since two channels existed, we used non-competing duplex reactions (two primer and probe pairs) to detect targets. However, there also exists other strategies to develop duplex assays including [14]; competing duplex reactions (one primer pair with two probes binding the same region); non-competing (hybrid) duplex reactions; and duplexing using non-specific double stranded DNA binding dyes/non-competing hybrid probe assays [15]. For the higher order multiplex assays, we developed the triplex probe mix assay (probe mix multiplexing) and the fourplex assay (amplitude based multiplexing).

Amplitude based multiplex assay design is quite similar to that of duplex double stranded DNA binding dye but with more targets (≥3). Essentially, since there are two channels, each channel can be used to detect two targets (T1-T4) at 0.5× and 1× concentrations of primers and probes as seen in Figure 8C. T1 and T2 are detected in Channel 1 (FAM) at 0.5× and 1× concentrations of primers and probes, respectively, while T3 and T4 are detected in Channel 2 (HEX/VIC) at 0.5× and 1× concentrations of primers and probes, respectively, giving rise to 16 (2^4^) droplet clusters in the 2D amplitude. In our work, we used four SARS-CoV-2 primer and probe pairs targeting RBD2, N, ORF1ab, and RPP30 to demonstrate this phenomenon. T1 and T2 were correspondent to RBD2 and N targets respectively, while T3 and T4 were corresponding to RPP30 and ORF1ab respectively. The separation of 16 clusters was achieved and additionally the exclusion of different targets using our sample matrix yielded expected results. Currently, no SARS-CoV-2 assay has been published or commercialized that demonstrate this approach.

Triplex SARS-CoV-2 ddPCR assays including two that use the ddPCR triplex probe mix assay approach have been developed by different companies for commercial purposes. Notably, these assays can be used in specific instruments and have different targets hence pose a challenge when one wants to change targets for research or clinical applications. To curb this, we used primers targeting ORF1ab, N, and RPP30 to demonstrate how to develop similar assays. Using the triplex probe mix approach, 8 droplet clusters are expected to arise from three targets (T1-T3) as seen in Figure 8C. In the 8 clusters, two targets T1 and T2 are detected in a conventional manner (100% probe concentration) in channel 1 (FAM) and Channel 2 (HEX/VIC) respectively. However, the third target (T3) is conjugated with half probe concentrations i.e. 50% FAM and 50% HEX/VIC to constitute the final 100% probe concentration hence located between T1 and T2 in the 2D amplitude with a consequent lowered amplitude in both Channels 1 and 2 amplitudes. Since the triplex assay was able to accommodate the recommended China CDC primers, it was used to show steps on optimization of multiplex assays.

After optimization tests, we analyzed the fitness of the triplex probe mix assay in detecting clinical samples and performing research. From our results, the assay was fit for SARS-CoV-2 diagnosis and research. Similar to the other ddPCR results [7,8,10,12,13], we also found that the N gene was more sensitive in detection of COVID-19 from low concentrated samples than the ORF1ab gene. This improved diagnostic sensitivity has been previously speculated to be due to the relative abundance of the N gene subgenomic mRNA produced during viral replication [21–23]. Since the triplex probe mix assay was fit for diagnosis, it could be used to greatly cut on costs when performing simplex assays for each target. Remdesivir has been extensively used in clinical applications after the drug was found to inhibit SARS-CoV-2 *in vitro* [24]. We used this drug and another drug we termed Code 30 to show the applicability of multiplex assays in performing research. As expected, the multiplex assay could quantify the activity of the two drugs against SARS-CoV-2. This may support the suggestion by Yu et.al to use ddPCR in quantifying antiviral efficacy against SARS-CoV-2 [13]. Only two non-diagnostic research applications of ddPCR against SARS-CoV-2 exist [9,11]. In both research, no multiplex assay was described as much as both used two primers targeting the ORF1ab and N gene. This leaves room for exploring the merits of multiplexing to perform research. We believe that multiplex assays can also help in testing how different targets respond when performing research like gene expression, copy number variations, and rare mutation detections.

## Conclusion

Primers and probes for developing assays capable of detecting SARS-CoV-2 already exists. Developing multiplex ddPCR assays using these resources will help to save on costs, increase sample throughput, and maximize on the number of targets that can be sensitively detected within a small sample. Here, we show steps on how to develop higher order multiplex assays that are fit for diagnosis of SARS-CoV-2 and also performing research related experiments for SARS-CoV-2. Prospective scientists are welcome to use our assay or develop similar assays depending on their targets to help in fight against SARS-CoV-2 and its associated disease COVID-19.

## Data Availability

No external data was used. All data is included in the manuscript

## Acknowledgements

All the researchers acknowledge the help and support provided by all students and staff at the Key Laboratory of Special Pathogens and Biosafety, Center for Biosafety Mega-Science, Wuhan Institute of Virology, Chinese Academy of Sciences. We also appreciate the University of Chinese Academy of Science and the Wuhan Institute of Virology, CAS, for facilitating and enabling us to conduct research under the “CAS Belt and Road Masters Fellowship and ANSO Scholarship for Young Talents”.

