## Supplementary material for "Developing multiplex ddPCR assays for SARS-CoV-2 detection based on probe mix and amplitude based multiplexing": Figures S1 S2 and Table S1

### Supplementary Information

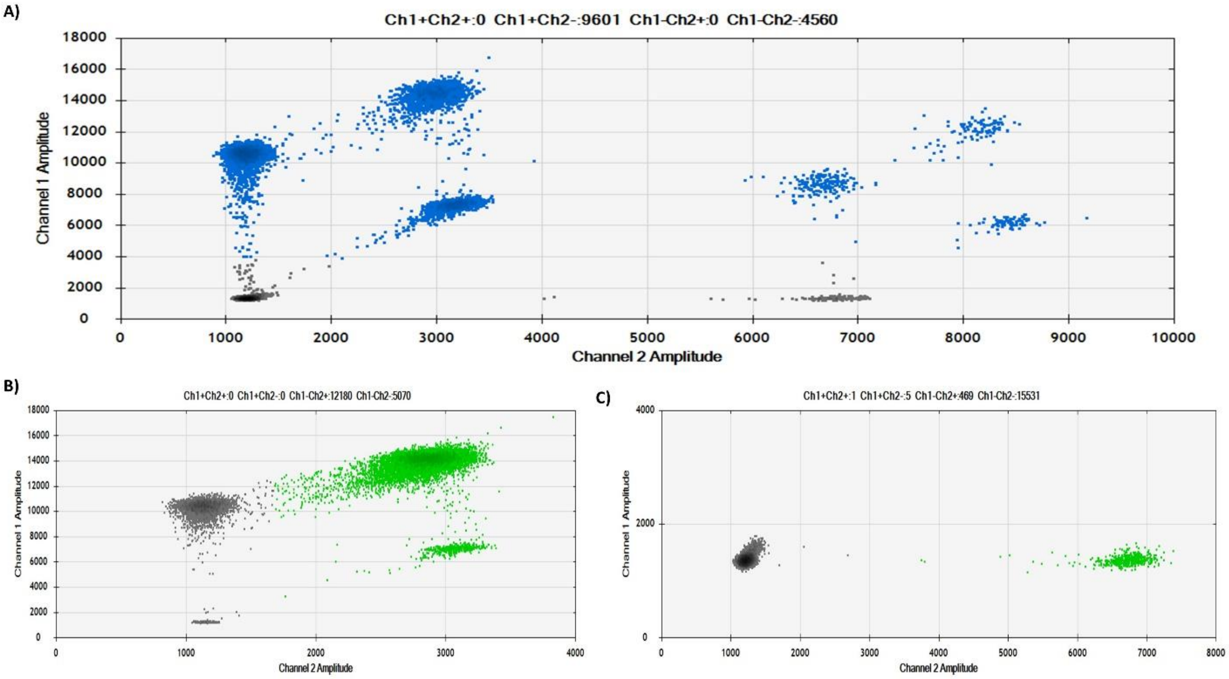


**Figure S1:** 2D amplitude QuantaSoft™ Software result from a triplex probe assay. A) Assay result with a sample spiked in a background of pooled human gene. B) Assay result of a virus only sample. C) Assay result of a sample containing only pooled human gene.

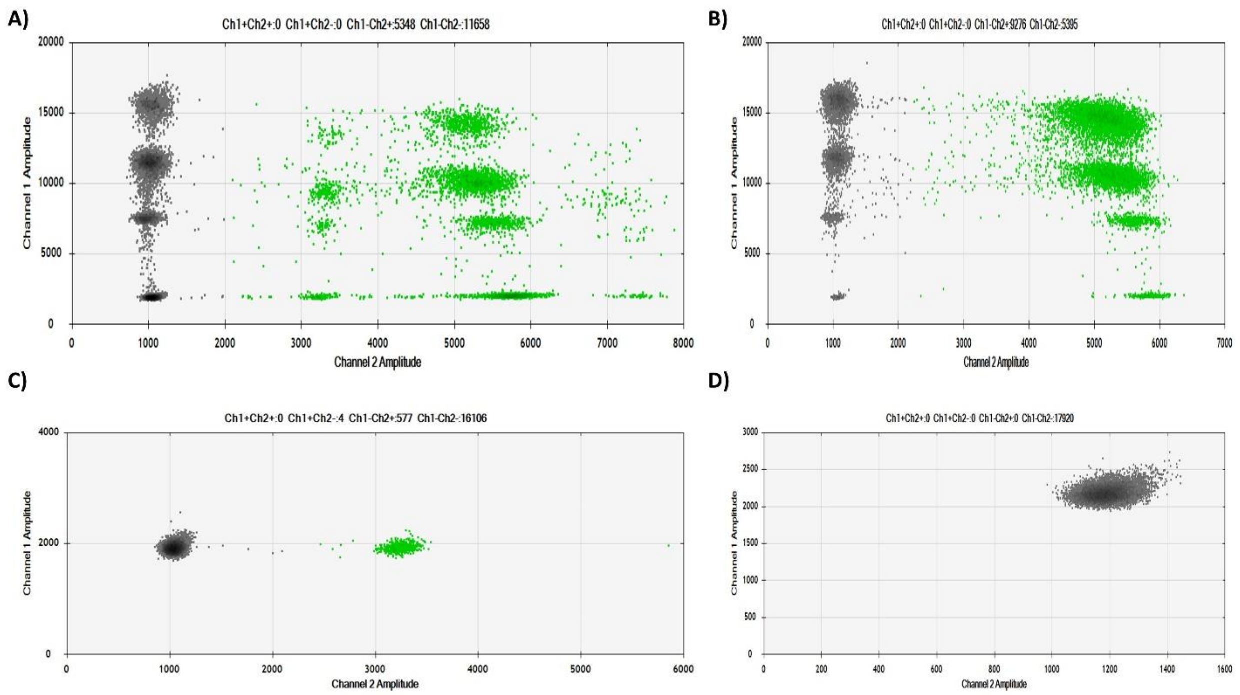


**Figure S2:** Droplet separation using a fourplex assay. A) A) SARS-CoV-2 spiked in a background of pooled human sample. B) SARS-CoV-2 sample. C) Pooled human sample. D) Negative sample.

### LoB calculations

**Table S1:** Limit of blank (LoB) results

| Sample | Total droplets | N Pos. droplets | ORF1ab Pos. droplets | IC (copies/µl) |
|---|---|---|---|---|
| 1 | 18206 | 0 | 0 | 426 |
| 2 | 18944 | 0 | 0 | 3.11 |
| 3 | 19991 | 0 | 0 | 52.5 |
| 4 | 19283 | 0 | 0 | 26.3 |
| 5 | 18781 | 0 | 0 | 1.25 |
| 6 | 19326 | 0 | 0 | 3.65 |
| 7 | 19982 | 0 | 0 | 15.9 |
| 8 | 19760 | 0 | 0 | 1440 |
| 9 | 18391 | 0 | 0 | 2010 |
| 10 | 19973 | 0 | 0 | 21.8 |
| 11 | 19104 | 0 | 0 | 0.0616 |
| 12 | 19223 | 1 | 0 | 1.84 |
| 13 | 18353 | 0 | 0 | 69.4 |
| 14 | 19968 | 0 | 0 | 3.54 |
| 15 | 19654 | 0 | 0 | 543 |
| 16 | 18722 | 0 | 0 | 28.3 |
| 17 | 18777 | 0 | 0 | 829 |
| 18 | 18685 | 0 | 0 | 53.6 |
| 19 | 17155 | 0 | 0 | 1460 |
| 20 | 19483 | 1 | 0 | 10.3 |
| 21 | 18893 | 1 | 0 | 18.1 |
| NTC | 19443 | 0 | 0 | 0 |
| NTC | 19143 | 1 | 0 | 0 |
| NTC | 19015 | 0 | 0 | 0 |
